# Laboratory tests for influenza and the respiratory syncytial virus may be used as a proxy for associated hospitalizations in Canada

**DOI:** 10.1101/2024.05.23.24307800

**Authors:** David Champredon

## Abstract

In Canada, the national number of hospital admissions associated with influenza and respiratory syncytial virus (RSV) is usually available about three months after admission. This delay hampers real-time analysis involving hospitalization data, like, for example, epidemic forecasting. Here, using a mixed-effects model on 15 years of data covering about 70% of the Canadian population, we show that these hospitalizations can be approximated using the number of laboratory tests positive for influenza A and RSV, a data stream publicly reported much more rapidly, about two weeks after symptoms onset.

## Introduction

Historically and before the emergence of COVID-19, seasonal influenza and the respiratory syncytial virus (RSV) are the most burdensome seasonal respiratory diseases on Canadian health systems. The ad-hoc database to extract hospitalizations associated with these two viruses across Canada is the Discharge Abstract Database (DAD), which captures administrative, clinical and demographic information on hospital discharges[1]. However, data availability is typically delayed by about three months because of the necessary data cleaning and quality assurance processes. This long reporting delay hampered efforts to analyze hospitalizations associated with respiratory diseases in or near real-time.

Another data source is available to monitor the spread of seasonal influenza and RSV (and other respiratory infections) on the public website of the Public Health Agency of Canada (PHAC) through its weekly respiratory virus detection reports[2]. This data set reports every week, with a two-week delay, the number and results of laboratory polymerase chain reaction (PCR) tests for multiple respiratory infections, including influenza and RSV. This relatively short delay from the actual time of infection makes this data more amenable to near real-time analysis of influenza or RSV infections across Canada (for example, forecasting the epidemic trajectory).

This study aims to assess if the data stream of laboratory tests performed for seasonal influenza and RSV can be used as a proxy for hospitalizations associated with these infections. We hypothesize that a steady relationship exists between hospitalizations and tests because, in Canada, most of the tests are performed in hospital settings.

## Methods

### Data

Hospital admissions data were retrieved from the DAD. We filtered the data for admission dates from September 1st, 2008, to September 1st, 2022. Then, we restrained our analysis to the most populous Canadian provinces and filtered data from the following provinces: British Columbia (BC), Alberta (AB), Saskatchewan (SK), Manitoba (MB) and Ontario (ON). We selected admissions from acute care facilities, nursing homes and emergency departments.

We defined an RSV-associated hospital admission as a patient whose had any of its 25 diagnostic codes contained at least one of the ICD-10 codes defining RSV, that is, J12.1, J20.5, J21.0, B97.4. Similarly, seasonal influenza-associated hospital admissions were defined based on the ICD-10 codes J09, J10, J10.0, J10.1, J10.8, J11, J11.0, J11.1, J11.8.

The laboratory test data (total volume of tests performed and the number of positive tests) was retrieved from the PHAC public website[2] from September 1st, 2008 to September 1st, 2022. Test data for influenza are typed, but not the hospitalization data. For reasons detailed in the Results section, we considered influenza A only and (in supplementary material) both influenza A and B. In the latter case, we summed the number of tests for both influenza A and B to compare test and hospitalization data for influenza. RSV data are not typed for both hospitalization and test data. Test data are reported as the aggregate number of tests and positives during one week.

We defined a season as starting on week 35 of the year and ending on week 34 of the following year. The hospitalization and test data are matched for the same week of the season (for a given province and virus). Hence, we implicit assume that a hospital admission and the report of the associated positive test result happen within the same week.

### Statistical models

We considered the log-transformed weekly number of hospitalizations and positive tests shifted by one (the shift of one is to avoid singularities when no positive test or hospitalization occurs in a week). We set *H*(*i, s, p, v*) = log(*h*(*i, s, p, v*) + 1) where *h*(*i, s, p, v*) is the number of hospitalizations associated with virus *v* during week *i* of season *s* in province *p*. Similarly, we have *T* (*i, s, p, v*) = log(*t*(*i, s, p, v*) + 1) the number of tests positive for virus *v* during week *i* of season *s* in province *p*. The motivation for log transforming the data is to fit equally well the hospital admissions data at all scales (a fit on the linear scale tends to overweight large values of the data). Moreover, when plotting the data before the statistical analysis, the relationship appears more linear on the log scale than on the untransformed scale.

#### Mixed effects model

If we expect similar relationships across provinces and seasons, we can use an inference framework that “shares information” between provinces and seasons. Hence, we consider a mixed-effects model to make a more global inference thanks to the implicit hierarchical structure of this type of model. We assume that the season and province variables are random effects, and consider the virus (influenza or RSV) a fixed-effect categorical variable. This mixed-effects model was implemented with the R package `lme4`[3] and was coded as: `lmer(H ∼ T*virus + (T|season) + (T|prov))`.

#### Linear regressions

We also consider simple linear regression models, independent from one another, to estimate the association between hospitalizations and positive tests for a given province, virus and season. This model was implemented in the R software as `lm(H ∼ T)`.

The inferences provided by these simple linear regressions should be considered only if we assume that each system consisting of the season, province, and virus triplet are independent of one another and that no additional information can be “borrowed” outside the focal season, province and virus.

### Leave-one-out validation

An important point for this analysis is demonstrate the possibility of performing a statistical regression to extrapolate hospital admissions from laboratory tests for a *new* season. Hence, we perform a leave-one-out validation by fitting the mixed-effects model to the entire data set except one season and then calculate the predicted hospital admissions using the observed number of laboratory tests that season. The quality of the prediction is assessed by calculating the proportion of *observed* hospital admissions (during the left-out season) that are within the 95% confidence interval of the *predicted* hospital admissions. The predictions are calculated using the `predict()` function on the output of the `lmer` object, and the confidence intervals are calculated using the function `predicInterval()` from the `merTools` R package[4] (the R code is provided as supplementary material). This leave-one-out validation is done on each one of the 15 seasons.

### Data availability

The laboratory test data is publicly available on PHAC’s public website and is also provided as a supplementary file (`data_share_labpos.csv`). The hospital admission data can be obtained by sending a request to the Canadian Institute for Health Information (CIHI).

### Hospital data perturbation

We do not have the permission to share the hospitalization data. However, to promote the reproducibility of this study, we provide a perturbed data set of hospital admissions in the supplementary file `data_share_hosp.csv` that was generated by adding random values to the original data set while ensuring the mean, variance and correlations of the perturbed data set are comparable to the original one.

Let *H*_private_ be the real number of hospital admissions for a given virus, province and week. The associated number shared in the supplementary file `data_share_hosp.csv`, *H*_shared_, is defined as *H*_shared_ = *X* + *Y*, where *X* is a normal distribution, rounded to the nearest integer, with mean *H*_private_ and a coefficient of variation equal to 0.2, and *Y* is generated from a Bernoulli distribution with probability 0.5. The shared data set resulting from this added random noise conserves the basic statistical properties (mean and between-group correlation, variance is larger because of the added noise) of the real–and private–data set, as shown in Figure S2.

The computer code, in the R programming language, implementing the full statistical analysis presented here is given in supplementary files `main_to_share.R`, `plot.R`, `stat.R` and `prediction.R`.

### Ethics approval and consent to participate

The Health Canada-Public Health Agency of Canada Research Ethics Board considered this study to be exempt from the requirement for research ethics review pursuant to article 2.2 of the Tri-Council Policy Statement: Ethical Conduct for Research Involving Humans.

## Results

### Data visualization

The paired data of the log number of hospital admissions (*H*) and log number of positive tests (*T*) is shown in Figure S1. The same plot is presented in Figure 1 when laboratory test results for influenza B are excluded. The linear relationship between hospitalizations and tests during most seasons across the selected provinces is highlighted in this figure by showing the linear regression for each season, province and virus (solid coloured line).

**Figure 1:**
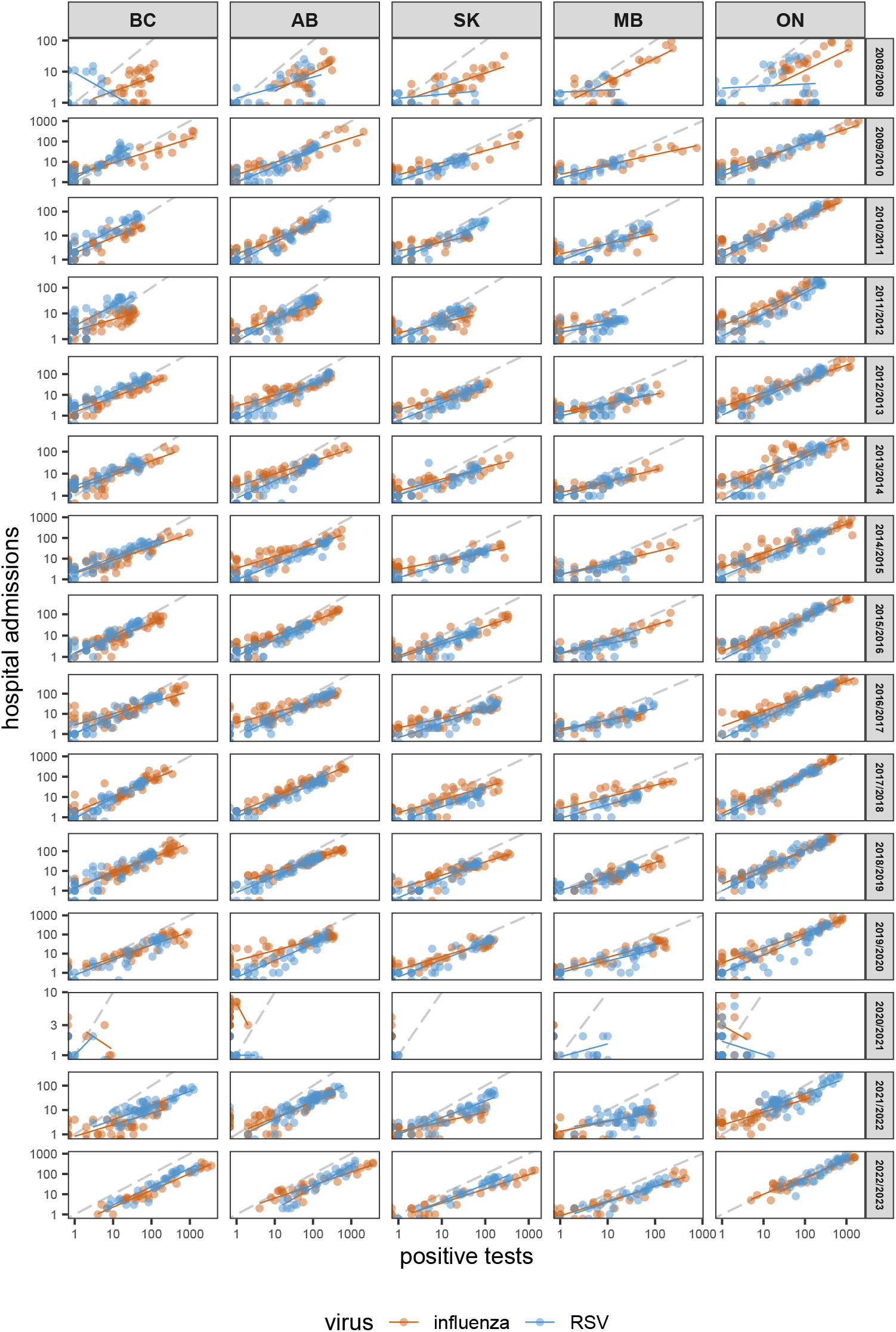
Hospitalization and test data. Each point corresponds to the number of hospital admissions and number of positive tests paired for the same week by virus, province and season. The solid colored line shows the simple linear regression of the data subset by virus, province and season. The virus labelled “influenza” includes results for laboratory tests for influenza type A only. The dashed grey line indicates the identity line when the intercept is 0 and the slope is 1. The data is shown using the logarithmic scale. Due to confidentiality restrictions, we are unable to share the original hospital admission data. Therefore, the perturbed version of the data is shown in this figure to protect privacy.

In the Northern Hemisphere, and particularly in Canada, influenza B does not consistently circulate every season, being barely present some seasons [5]. Comparing Figure 1 and Figure S1, we see that including laboratory results for influenza B apparently affects the log-linear relationship with hospitalizations. Hence, for the primary analysis in this study, the data set for the influenza laboratory test results *excludes* influenza type B. Only test results for influenza A and RSV are used in the results presented below. The main analysis including influenza B test results is presented in supplementary material (Figure S9, Figure S10).

### Mixed-effects model

The mean estimates of the random coefficients (intercept and slope) across seasons and provinces groupings are shown in Figure 2. The values of all mean estimates are reported in Table S1 and the random effects (that capture the variability not explained by the fixed effects and are specific to the individual group) are shown in Figure S3.

**Figure 2:**
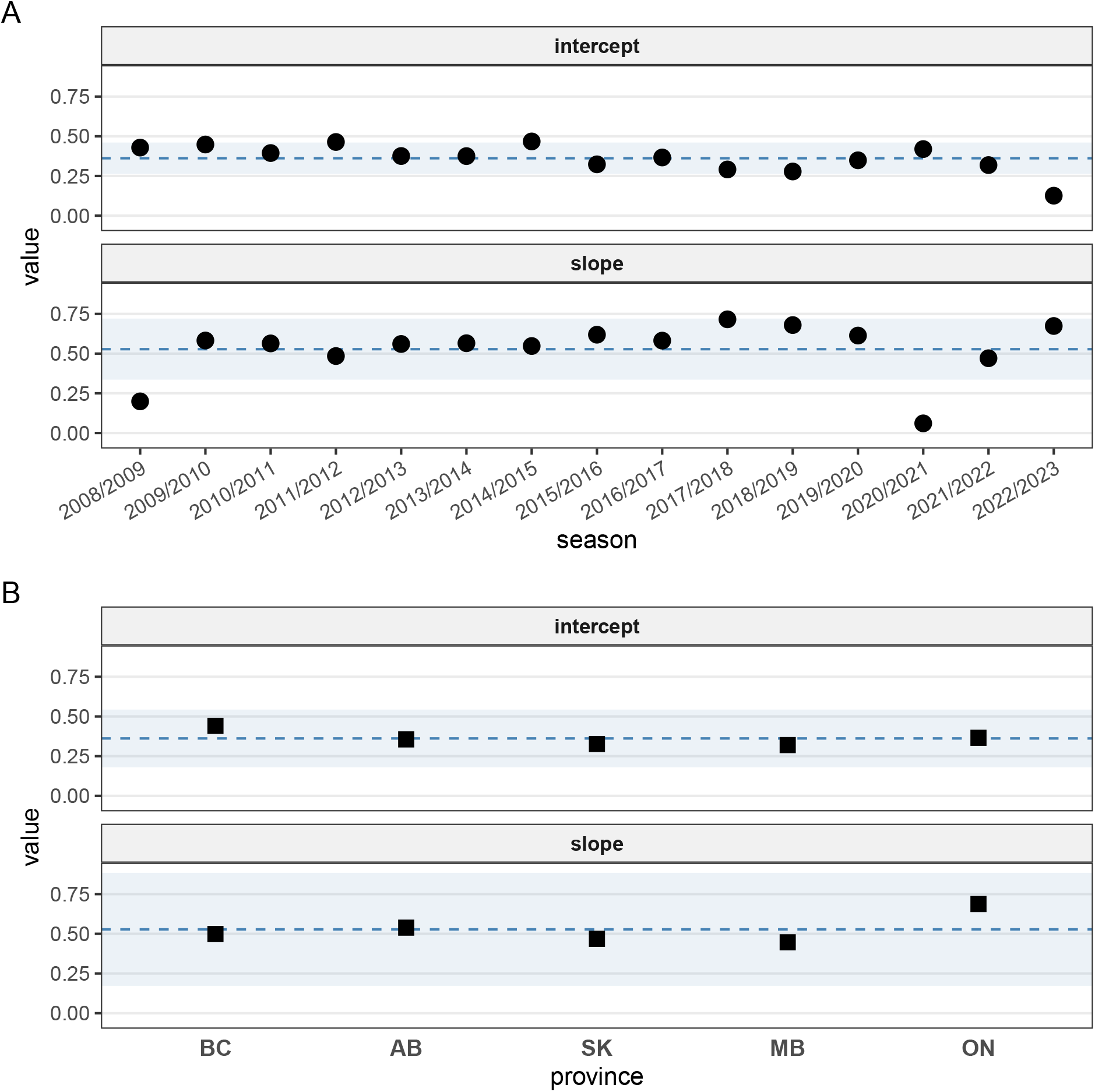
Mean estimates of the mixed-effects model. Mean estimates of the random coefficients (intercept and slope) of the mixed-effects model for the seasons group (panel A) and the provinces group (panel B). The horizontal dashed line represents the mean estimates of the associated fixed effect, and the horizontal light ribbon shows the width of +/- 2 standard deviations of the fixed effect. See Figure S3 for the random effects conditional values and variances.

The mean estimates of the intercept at the season level have similar values bounded between approximately 0.205 and 0.395, except for the 2022/2023 season with an estimate of 0.056. Likewise, the mean estimates for the slope by seasons are between 0.518 and 0.753, except for seasons 2008/2009 and 2020/2021 that have means estimates of 0.216 and -0.001 respectively (Figure 2, panel A and Table S1). The mean estimates of the slopes at the province level also have similar values between 0.494 and 0.563 for AB, BC, MB and SK. The mean level for ON is higher at 0.701 (Figure 2, panel B).

The estimates for the coefficients associated with the fixed effects from the virus variable (RSV versus the influenza reference) are -0.244 (95% CI: -0.264 ; -0.224) and 0.124 (95% CI: 0.108 ; 0.139).

The inferences above indicate that season-level estimates for 2008/2009 and 2020/2021 are markedly different from the other seasons. These seasons were impacted by two global pandemics: H1N1 for 2008/2009 (that occurred mainly during the summer of 2009 in Canada) and the SARS-CoV-2/COVID-19 for 2020/2021 (which is the season that had the most social distancing measures in Canada and reduced greatly the circulation of most respiratory viruses). To assess the impact of these “pandemic” seasons on the mixed-effects model estimates, we ran a sensitivity analysis that excluded these seasons. The comparison is shown in Figure S4 and shows a limited impact of including pandemic seasons on the inference.

### Independent linear regressions

The coefficients of the simple, independent, linear regressions on data subsets are shown in Figure S5. We note that, in general, for a given province and virus, there is moderate variability of the regression coefficients across “non-pandemic” seasons, that is all seasons except 2008/2009 (H1N1), 2020/2021, 2021/2022 and 2022/2023. This is highlighted in Figure S6, which provides a summary of the coefficients for the “non-pandemic” seasons. We also note that the *R*^2^ coefficient is often above 0.75, indicating a strong linear relationship between the logarithmic values of hospitalizations and positive tests.

We compare the estimates of these independent linear regressions with the coefficient estimates of the mixed-effects model in Figure S7.

### Leave-one-out predictions

The performance of leave-one-out predictions is shown in Figure 3 where the proportion of actual observations inside the predicted 95% confidence interval is plotted for each province and virus. Each boxplot is derived from 15 data points (one for each of the 15 seasons left out). For each season, the proportion is calculated from 52 (or 53 for leap years) data points, corresponding to the number of weeks in a season. The proportions of observations that lie within the 95% confidence interval of their predictions are mostly above 80% (Figure 3). The scatter plot comparing predictions with observations of hospital admissions is shown in Figure S8.

**Figure 3:**
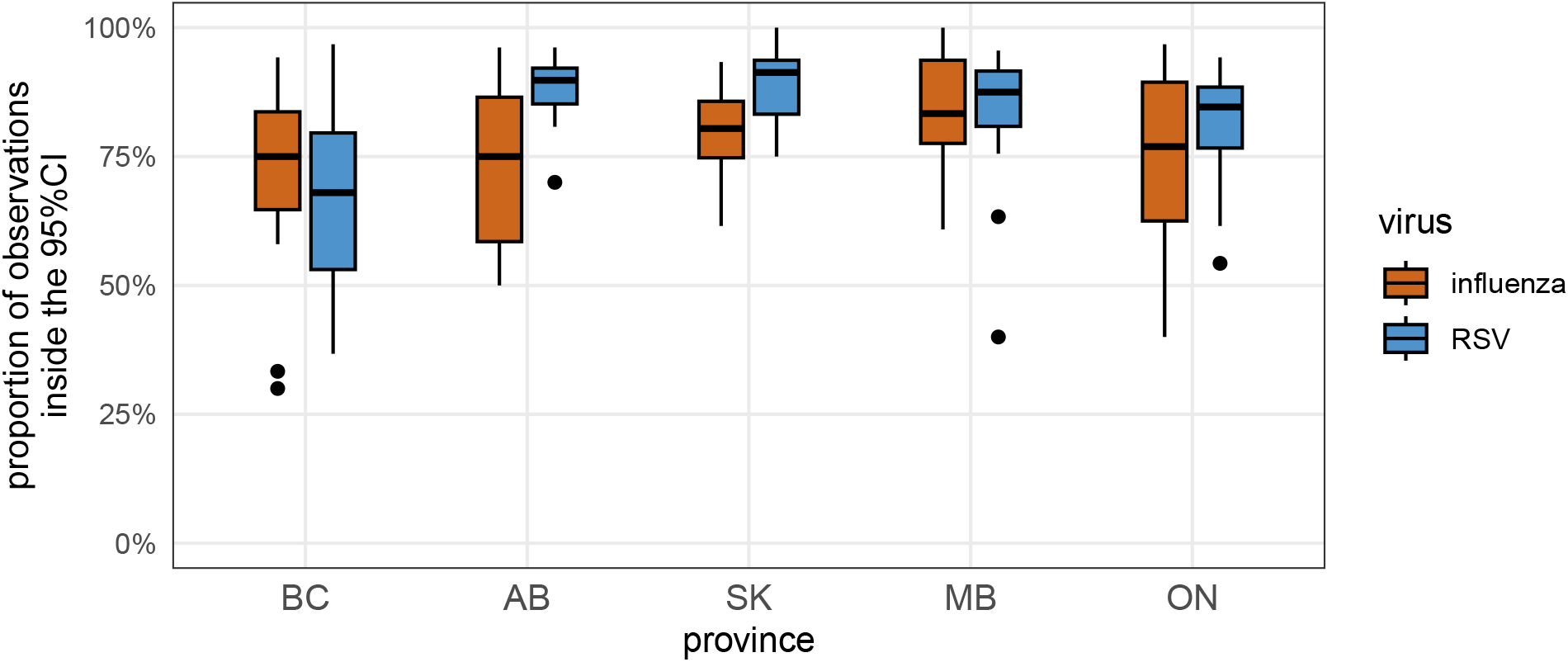
Leave-one-out predictions. Each boxplot summarizes the proportion of the observed hospital admissions that lie within the 95% confidence interval of the predicted hospital admissions for each season left out. The proportions are stratified by province and virus.

## Discussion

In Canada, the reports of hospitalizations associated with acute respiratory infections are available only about three months after the admission date. This relatively long delay can be detrimental to studies that need hospitalization data in near real-time (like, for example, epidemic forecasting). On the other hand, laboratory tests performed to detect acute respiratory infections are available within about two weeks after specimen collection. Moreover, those tests are usually done in hospital settings. Hence, this study focused on assessing if the number of positive laboratory tests associated with influenza A and RSV (available rapidly but not so relevant as an indicator of burden on health systems) could be a proxy for hospital admissions (lagging by several months but very relevant as a burden indicator) in Canada.

Using hospitalization and clinical surveillance data over 15 seasons (from 2008 to 2022), we fitted a mixed effects model and estimated a strong relationship between hospitalizations and positive tests for influenza A and RSV across five large Canadian provinces covering approximately 70% of the national population. The existence of such a relationship is not surprising because testing for influenza and RSV is mostly performed in healthcare settings in Canada. The added value of the present analysis is to quantify this relationship, assess how it varies in time (across 15 seasons) and in different jurisdictions (five provinces), and suggest a statistical approach to “nowcast” hospitalizations using ubiquitous laboratory test results.

Pandemics are global epidemiological events that usually trigger a strong public health response to reduce morbidity and mortality. The latest influenza pandemic emerged during the summer of 2009 and involved an A/H1N1 strain stemming from a triple reassortment of swine, avian and human flu viruses[6]. Given the perceived increased virulence of this strain when it emerged (because of its zoonotic reassortment)[7], this pandemic elicited a strong public health response globally, including in Canada, consisting of increased influenza testing. The COVID-19 pandemic had an even larger impact on health systems globally when it emerged in late 2019[8]. In particular, the stringent public health measures implemented in Canada during 2020 and 2021 had an unprecedented impact on the circulation of respiratory viruses and their surveillance[9]. These two pandemics clearly impacted the hospitalizations/tests relationship for influenza–as well as for RSV–as shown by our slope estimates: the mean slope was around 0.6 during non-pandemic years but dropped to about 0.1 during both pandemics (Figure 2 panel A and Table S1). In other words, during non-pandemic years, there are about 1.7 (=1/0.6) log tests performed for each log hospitalization, whereas this is multiplied nearly six times to 10 (=1/0.1) log tests during a season when a pandemic happens. Hence, during these two pandemics the standard, and relatively stable, hospitalizations/tests log-linear relationship was altered in most of Canada because testing for respiratory viruses surged.

Note that the intercept of the log-linear regression was not significantly affected by these pandemics (Figure 2 panel A and Table S1). Instead, we see a trend where the slope estimates seem to be decreasing since 2020/2021. A smaller slope means that fewer hospitalizations occur without performing a test. One possible explanation of this trend is the continuing increased surveillance of SARS-CoV-2/COVID-19 infections because of its high mutation rate, giving rise to multiple immune-escape variants[10]. This plausible heightened COVID-19 surveillance may cause more multiplex testing (including for influenza and RSV) than pre-COVID times in hospitals. Future data may confirm this trend and, were it to continue, if the testing efficiency would be negatively impacted (this would be reflected in a negative estimate for the intercept, meaning there are too many tests performed for the number of hospitalizations).

The analysis at the province level highlighted a small difference between the province of Ontario and the four other provinces. While intercept estimates of the five provinces have similar values, the slope estimate for the province of Ontario is larger (0.701) than the four other provinces (see Figure 2 panel B and Table S2). The interpretation of this result likely requires an in-depth analysis that is beyond the scope of this study and might be tied to different testing and/or hospitalization practices potentially driven by different socioeconomic environments.

An important insight of this study is to justify the use of surveillance laboratory test data as a proxy for hospital admissions for influenza and RSV in each province. Indeed, in Canada, the latter is available with at least a 3-month delay, whereas the former has about a 2-week delay. Hospitalizations are usually regarded as a more relevant metric than tests when considering the burden of respiratory infections on the health system. The long time lag in obtaining the hospitalization data hampers real-time analyses (*e*.*g*., forecasting incidence trajectories for seasonal influenza and RSV). Here, we showed that the weekly log number of tests could be used to approximate the weekly log number of hospitalizations associated with influenza and RSV in five Canadian provinces.

We also showed the mixed effects model may be used to generate hospital admissions proxy data (from laboratory test data) for a currently unfolding influenza and/or RSV season because it had a satisfactory accuracy on historical data (as measured by the metric shown in Figure 3). At the start of the new season, with little to no test data available, the estimates for the intercept and slope of the linear relationship will be close (or equal, if no data is available yet) to the associated fixed-effects model. As new laboratory test data come in, the estimates of the regression coefficients will incorporate this new information to provide, hopefully, more accurate estimates for hospital admissions.

Note that using the mixed-effects framework implies that the investigator assumes that incorporating information from the surveillance data of other provinces is relevant (say, because their health systems–as far as respiratory infections are concerned–are very similar). However, should the investigator assume that their jurisdiction of interest is unique, then an independent simple linear regression (as presented as an alternative modelling analysis in this study; see Figure S5 and Figure S6) may be better adapted for their analysis.

The analysis presented here has several limitations. First, it uses Canadian data only; hence, any extrapolation to other jurisdictions in Canada not included in our fitting data set (*e*.*g*., province of Québec, any Canadian Territories), or outside of Canada, may not be appropriate. In particular, the exclusion of test results for influenza B may not be a good approach in regions that see a more consistent circulation of this virus over time. The suggestion made here to use this statistical framework to estimate influenza or RSV hospital admissions for laboratory test counts may not be valid even for the jurisdictions where the data was used to fit the model. Indeed, this relationship depends on testing and hospitalization policies. If these policies suddenly change for the province(s) of interest, inferences about hospitalizations may be incorrect as they will be made from a model that draws information from other provinces and past data.

In conclusion, this study suggests that for investigators needing recent influenza/RSV hospital admission data from Canada but needing timely access, it may be possible to use the more rapidly reported laboratory test data to calculate proxies for influenza/RSV hospital admissions. The statistical framework presented here may also be used with other jurisdictions having similar epidemiological data reporting timelines.

## Data Availability

The laboratory test data is publicly available on the Public Health Agency of Canada website (https://www.canada.ca/en/public-health/services/surveillance/respiratory-virus-detections-canada.html)
The hospital admission data can be obtained by sending a request to the Canadian Institute for Health Information (CIHI).

## Acknowledgments

I thank Drs. Devan Becker (Laurier University, Waterloo, ON, Canada), Benjamin Bolker and Jonathan Dushoff (McMaster University, Hamiton, ON, Canada) for insightful discussions and comments on the statistical methodology; Dr. Abbas Rahal (Public Health Agency of Canada, Ottawa, ON, Canada) for assistance with the SAS script to retrieve the DAD hospitalization data.

