## supplementary material for "Laboratory tests for influenza and the respiratory syncytial virus may be used as a proxy for associated hospitalizations in Canada"

### Supplementary Material
### Laboratory tests for influenza and RSV can be a proxy for associated hospitalizations in Canada

David Champredon

May 23, 2024

### Data including influenza B laboratory tests

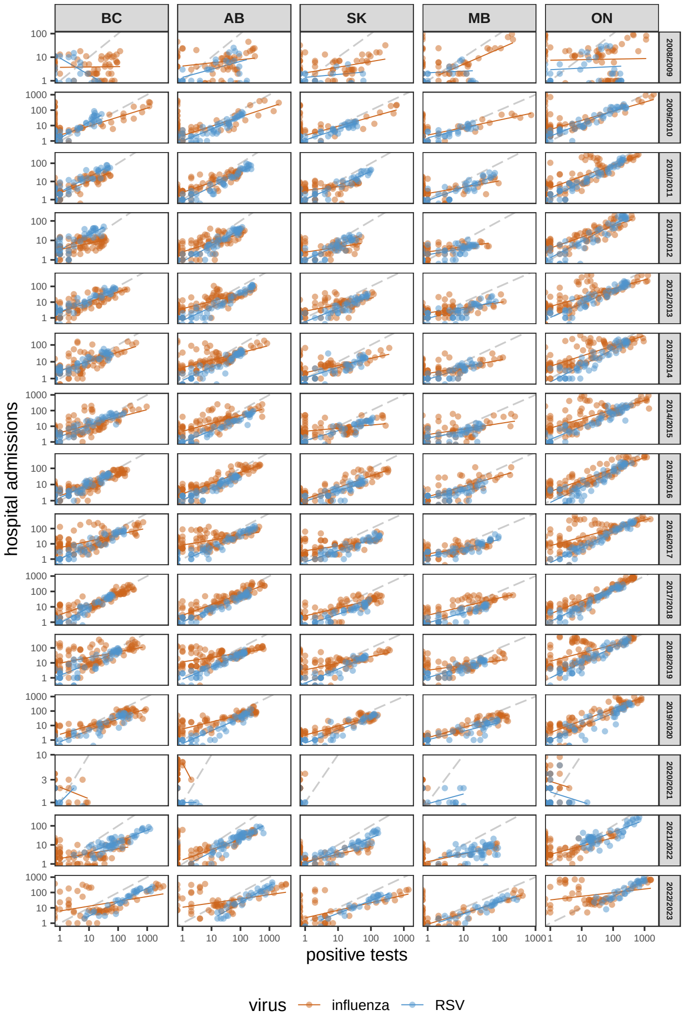


Figure S1: **Hospitalization and test data**. Each point corresponds to the number of hospital admissions and number of positive tests paired for the same week, by virus, province and season. The solid colored line shows the simple linear regression on the data subset by virus, province and season. The virus labelled "influenza" includes results for laboratory test for influenza for both types A and B. The dashed grey line indicates the identity line when the intercept is 0 and the slope is 1. The data is shown using the logarithmic scale.

### Table S1 - Mixed-effects model mean estimates at the season level

| | (Intercept) | logtests | virusRSV | logtests:virusRSV | season |
|---|---|---|---|---|---|
| 2008/2009 | 0.43 | 0.20 | -0.22 | 0.11 | 2008/2009 |
| 2009/2010 | 0.45 | 0.58 | -0.22 | 0.11 | 2009/2010 |
| 2010/2011 | 0.39 | 0.56 | -0.22 | 0.11 | 2010/2011 |
| 2011/2012 | 0.46 | 0.48 | -0.22 | 0.11 | 2011/2012 |
| 2012/2013 | 0.38 | 0.56 | -0.22 | 0.11 | 2012/2013 |
| 2013/2014 | 0.38 | 0.57 | -0.22 | 0.11 | 2013/2014 |
| 2014/2015 | 0.47 | 0.55 | -0.22 | 0.11 | 2014/2015 |
| 2015/2016 | 0.32 | 0.62 | -0.22 | 0.11 | 2015/2016 |
| 2016/2017 | 0.37 | 0.58 | -0.22 | 0.11 | 2016/2017 |
| 2017/2018 | 0.29 | 0.72 | -0.22 | 0.11 | 2017/2018 |
| 2018/2019 | 0.28 | 0.68 | -0.22 | 0.11 | 2018/2019 |
| 2019/2020 | 0.35 | 0.61 | -0.22 | 0.11 | 2019/2020 |
| 2020/2021 | 0.42 | 0.06 | -0.22 | 0.11 | 2020/2021 |
| 2021/2022 | 0.32 | 0.47 | -0.22 | 0.11 | 2021/2022 |
| 2022/2023 | 0.13 | 0.67 | -0.22 | 0.11 | 2022/2023 |

### Table S2 - Mixed-effects model mean estimates at the province level

| | (Intercept) | logtests | virusRSV | logtests:virusRSV | prov |
|---|---|---|---|---|---|
| BC | 0.44 | 0.50 | -0.22 | 0.11 | BC |
| AB | 0.36 | 0.54 | -0.22 | 0.11 | AB |
| SK | 0.33 | 0.47 | -0.22 | 0.11 | SK |
| MB | 0.32 | 0.45 | -0.22 | 0.11 | MB |
| ON | 0.37 | 0.69 | -0.22 | 0.11 | ON |

### Comparison between private and shared data

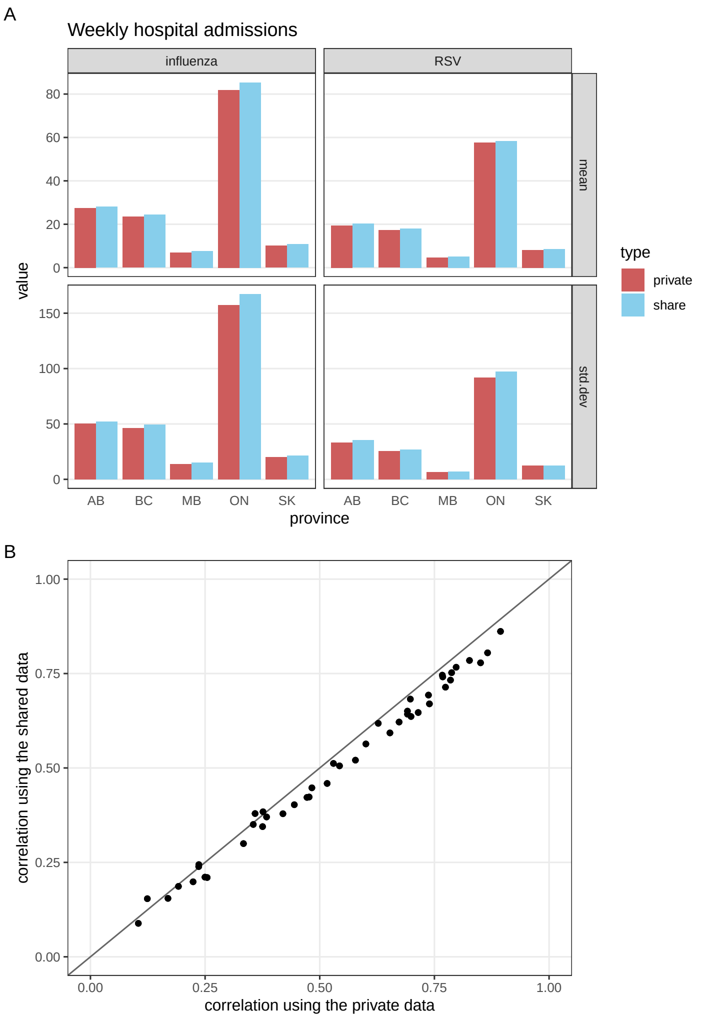


Figure S2: Comparison between the real number of hospital admissions and the shared data set. Panel A compares the mean and standard deviation of the real, and private, number of weekly hospital admissions and the perturbed data set shared with this study. The mean is calculated by province and virus across all seasons. Panel B compares the pairwise correlations by province and virus (*e.g.*, RSV in Ontario v.s. influenza in Manitoba) calculated using the private data set and the data set shared in this study. We note that the correlation is slightly lower for the shared data set as a result of the added random noise.

### Random effects of the mixed-effects model

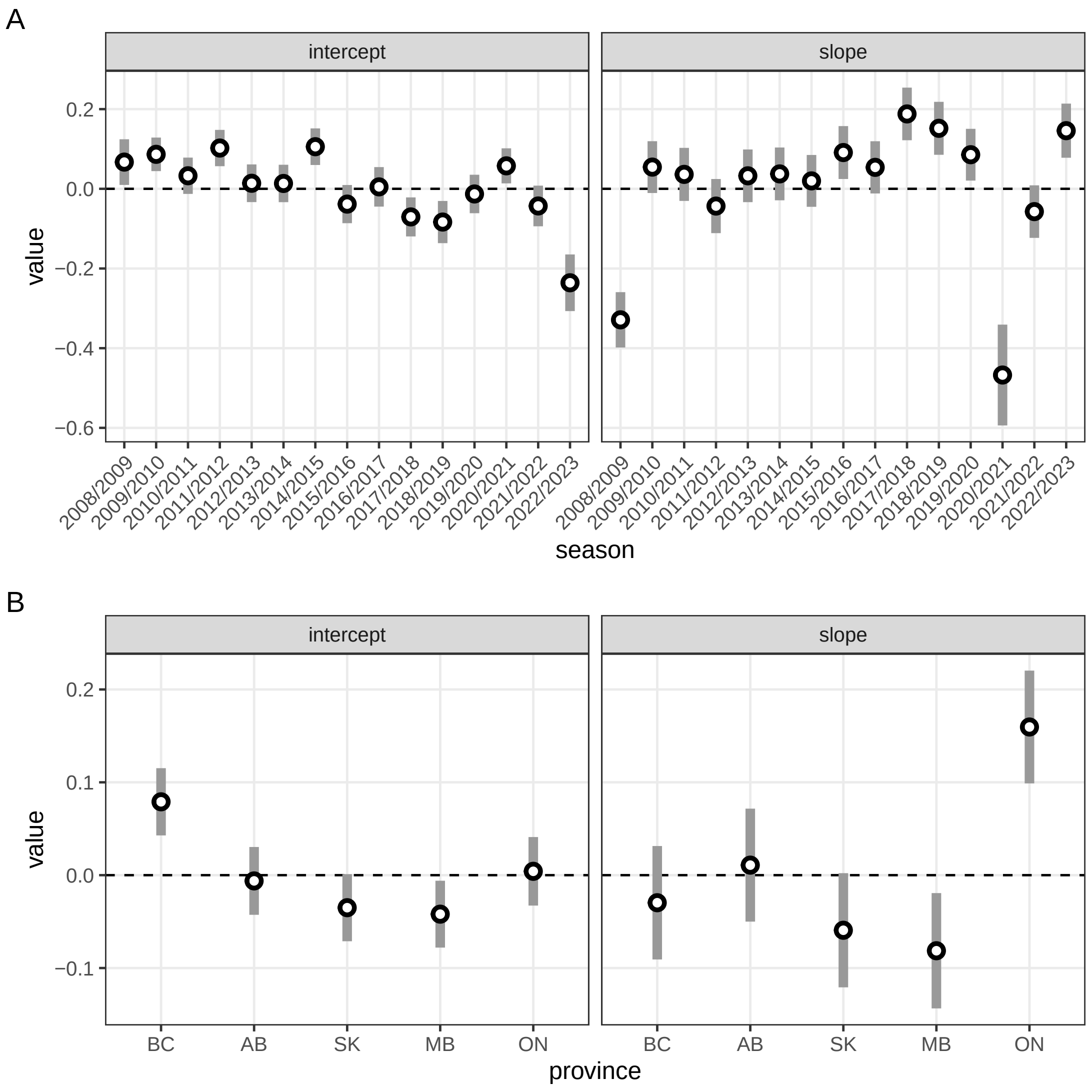


Figure S3: Random effects - The points represent the conditional mode for each group and level and the vertical segment 2 times the conditional standard deviation (as returned by the function `ranef()` of the `lme4()` package.) They capture the variability not explained by the fixed effects and are specific to the individual group. Panel A: random effects for the season group. Panel B: random effects for the grouping by province.

### Pandemic years exclusion

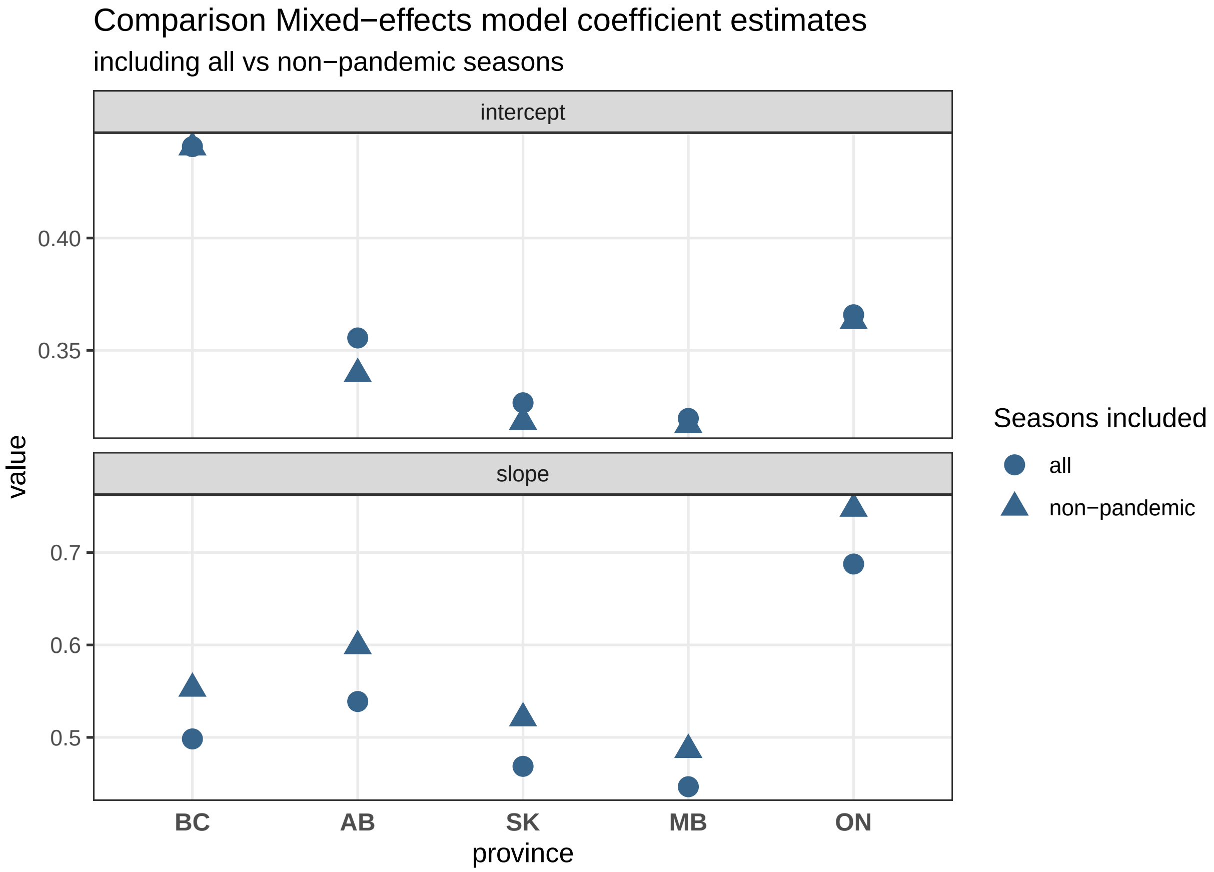


Figure S4: Impact of pandemic years on the mixed effects model - The triangle-shaped points show the mean estimates of the intercept (top panel) and slope (bottom panel) when the mixed-effects model is fitted on the data set that excludes the pandemic seasons (2008/2009 and 2020/2021). The circle-shaped points indicate the estimates when all 15 seasons are included in the fitting data set (as presented in the main manuscript).

### Estimates from the linear regressions

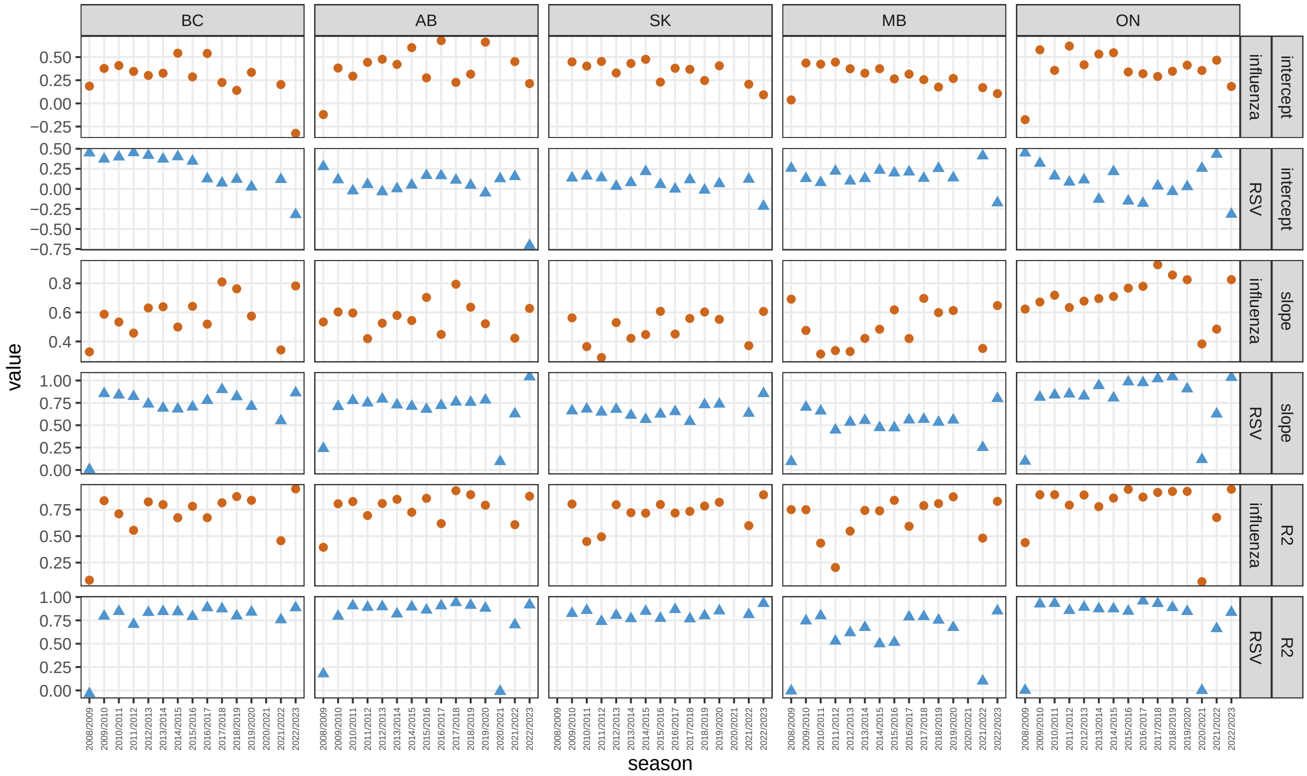


Figure S5: Coefficients from the linear regression between the logarithmic values of hospitalizations and positive tests. Each column represent a province and each row a coefficient type (intercept, slope, $R^2$) and a virus (influenza, RSV). The points represent the value of the regression coefficient for a given province, virus, season and coefficient type (to improve the clarity of the figure, their color and shape changes according to the virus (brown point for influenza, blue triangle for RSV).

13 # Summary of the estimates from the linear regressions

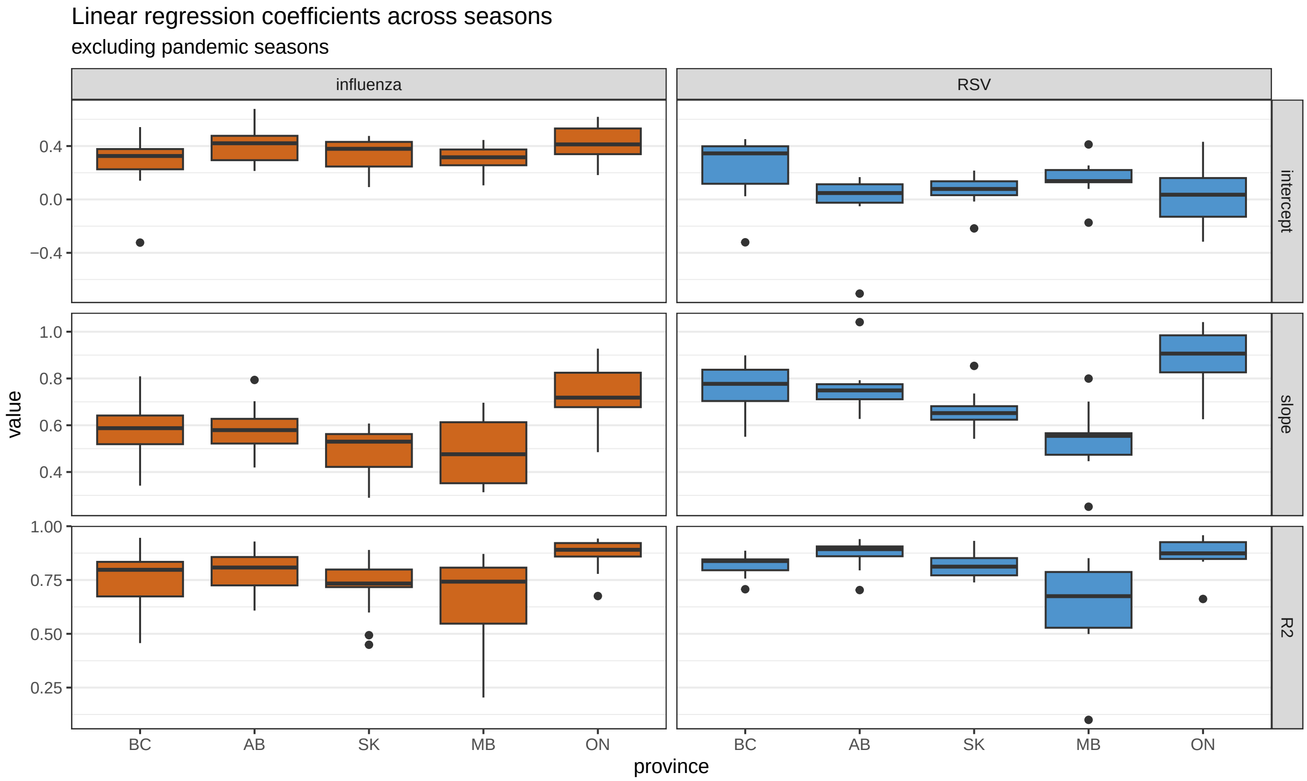


Figure S6: Summary of the coefficients from the linear regression between the logarithmic values of hospitalizations and positive tests.

### Comparison simple linear regression v.s. mixed effects model

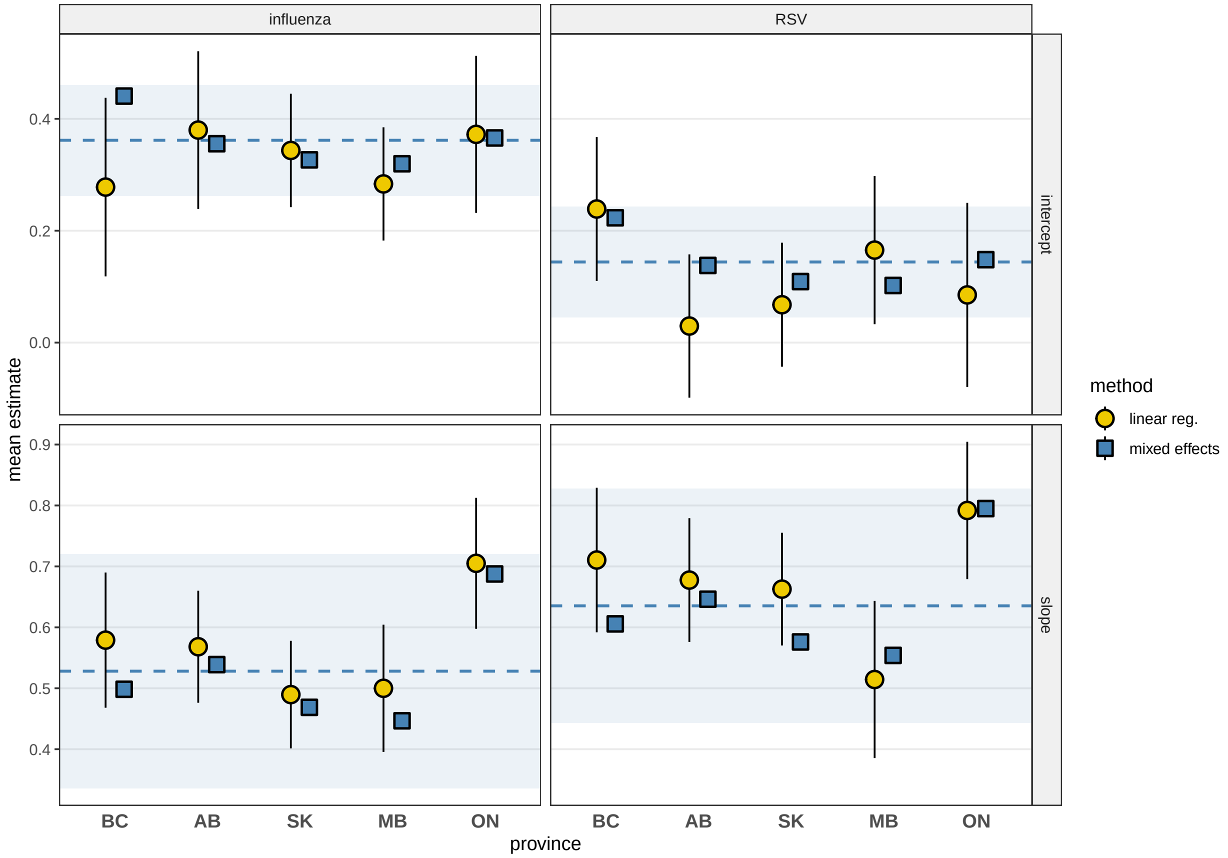


Figure S7: Inference comparison between a simple linear regression and a mixed-effects model. The circles represent the mean estimate when performing a simple linear regression on each single province (across all seasons) and the vertical error bar shows their 95%CI. The solid squares indicate the mean estimates from the mixed-effect models, the horizontal dashed line represents the estimate of its fixed effect and the light ribbon shows its +/-2 standard deviations (as in Figure 2 in the main text).

### Mixed-effects model predictions v.s. observations

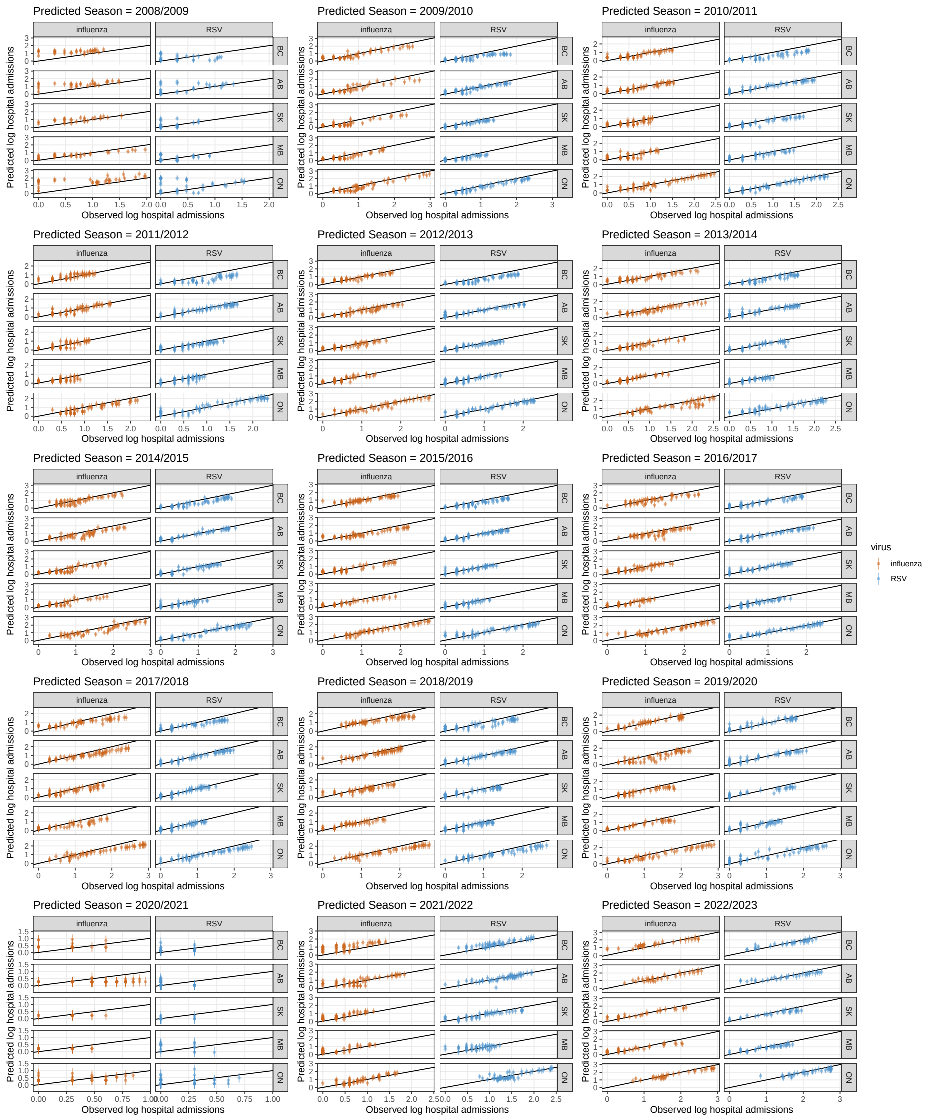


Figure S8: Comparison between the observed and predicted log number of hospital admissions. Each panel represent a predicted season (the one left out of the fitting data set). The points represent the mean estimate (from the mixed effects model) of the log number hospital admissions and the vertical segment its 95%CI. The solid black line is the bisect line (“y=x”) for a visual comparison with perfect predictions.

### Main results including influenza B laboratory tests

In this section, we present the same results as the ones presented in the main text, but including the weekly number of positive influenza B test results (the analysis shown in the main text uses influenza A only).

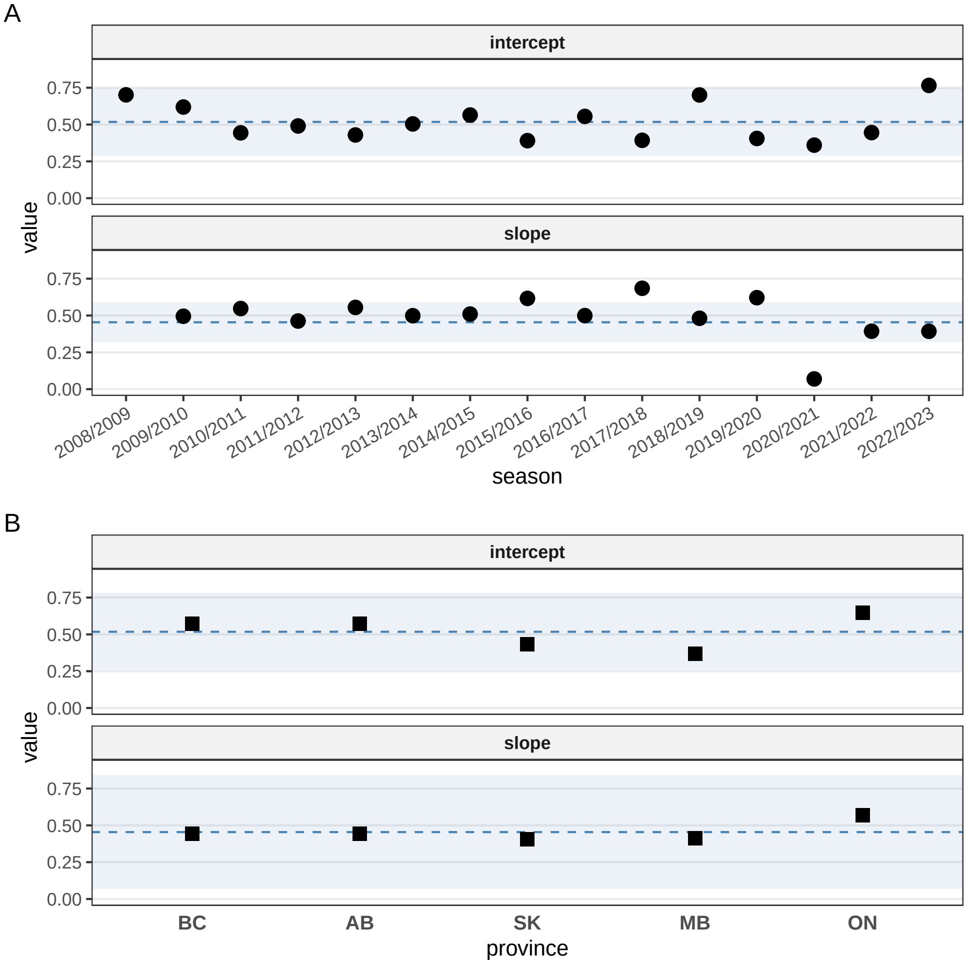


Figure S9: **Mean estimates of the mixed-effects model, including influenza B.** Mean estimates of the random coefficients (intercept and slope) of the mixed-effects model for the seasons group (panel A) and the provinces group (panel B). The horizontal dashed line represents the mean estimates of the associated fixed effect and the horizontal light ribbon shows the width of +/- 2 standard deviations of the fixed effect.

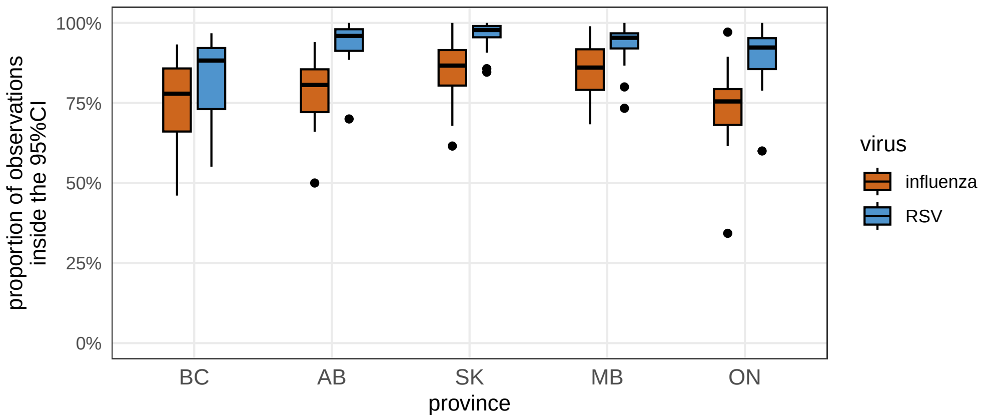


Figure S10: **Leave-one-out predictions, including influenza B.** Each boxplot summarizes the proportion of the observed hospital admissions that lie within the 95% confidence interval of the predicted hospital admissions for each season left out. The proportions are stratified by province and virus.

End of supplementary file.
